# Buccal DNA Methylation-Derived EpiScores are Associated with Neonatal Morbidity Burden in Infants born Very Preterm

**DOI:** 10.64898/2026.09.25.26363955

**Authors:** Priyadarshni Patel, Siyu Zhu, Neha Sehgal, Marie Camerota, Brian S. Carter, Jennifer Check, Jennifer B. Helderman, Julie A. Hofheimer, Jonathan S. Litt, Elisabeth C. McGowan, Charles R. Neal, Steven L. Pastyrnak, Lynne M. Smith, T. Michael O’Shea, Carmen Marsit, Barry Lester, Todd M. Everson

## Abstract

**Background:** Infants <30 weeks gestational age (very preterm (VPT)), are at risk of experiencing multiple serious neonatal morbidities. These morbidities impart serious biological stress, which may be reflected in dysregulated protein expression even after the morbidities have resolved. Recent studies suggest that DNA methylation-based predictors of circulating proteins may reflect the molecular effects of neonatal adversity. The objective of this study is to examine associations between DNA methylation-derived EpiScores, which are externally trained methylation signatures that proxy circulating protein levels, and neonatal morbidity burden in children born VPT. Buccal DNA methylation was obtained at NICU discharge in 537 infants from the NOVI cohort. A total of 111 EpiScores were calculated and tested for association with multimorbidity (0-1 vs. ≥2 morbidities) using adjusted linear regression models. Cross-tissue analyses in an external paired blood-buccal dataset (n=21) were used to identify EpiScores with moderate-to-strong correlations across tissues.

**Results:** Eighteen EpiScores were nominally associated with multimorbidity (p < 0.05), with Melanoma-derived growth regulatory protein (MIA) with FDR < 0.05 (β = 0.0023, p = 4.4 × 10□□). In cross-tissue analyses, 35 EpiScores yielded r > 0.50 (p<0.05), 11 of which showed nominal associations with multimorbidity, including MIA. Overall, the morbidity-associated EpiScores reflected immune, inflammatory, metabolic, and growth-related pathways.

**Conclusions:** Buccal DNA methylation-derived EpiScores capture biological signatures associated with neonatal morbidity burden in very preterm infants.

## Background

Infants born <30 weeks gestational age (GA) face a high risk of neonatal morbidities, including bronchopulmonary dysplasia (BPD)(1), severe retinopathy of prematurity (ROP) (^2^), necrotizing enterocolitis (NEC)(3), sepsis (4), and brain injury(5). These complications frequently co-occur and contribute to long-term neurodevelopmental (6), respiratory, and metabolic impairments that extend into childhood and adulthood(7). Our previous work has demonstrated that a greater number of neonatal morbidities may perturb biological processes important for growth and development (8), and are associated with early childhood cognitive and motor impairments (9). Similarly, others have shown that cumulative neonatal morbidities are associated with extended NICU stays (10), and with neurodevelopmental impairments in childhood (11–14). Despite the clinical consequences of neonatal multimorbidity, how cumulative morbidity burden shapes persistent molecular alterations remains poorly understood.

Inflammation, immune dysregulation, and altered metabolic signaling play central roles in the pathogenesis of preterm complications(15, 16). Very preterm infants are exposed to various infections, oxidative stress, and medical interventions in the neonatal intensive care unit (NICU)(17). These exposures may induce changes in DNA methylation that alter gene regulation and downstream biological pathways involved in disease development. We have demonstrated that preterm-related multi-morbidity is associated with differential DNA methylation (8), and with accelerated epigenetic aging(18, 19), suggesting that epigenetically derived indicators can provide insight into neonatal health.

DNA methylation derived epigenetic scores (EpiScores) provide a novel and promising approach to capture complex biological processes using composite DNA methylation patterns that proxy circulating proteins(20). EpiScores are externally trained DNA methylation signatures developed in large adult cohorts using machine learning approaches that predict circulating protein concentrations from blood-based methylation patterns. Rather than directly measuring neonatal protein levels, EpiScores act as epigenetic proxies for protein-related biological pathways and have been externally validated across multiple adult cohorts. Through this framework, multiple EpiScores have undergone external validation, demonstrating robust performance across independent cohorts(20) (21). The proteins captured by these validated scores are involved in a wide range of biological processes, including inflammation, immune response, metabolic regulation, and cardiovascular function(20). This approach provides a tool to leverage DNA methylation data to estimate and study how protein activities are related to health characteristics. However, their application to neonatal populations, particularly very preterm infants, and their relevance to early-life morbidity burden remain largely unexplored.

Buccal epithelial cells offer a non-invasive and developmentally relevant tissue for epigenetic profiling in neonates, particularly when blood collection is limited(22). However, EpiScores were developed using blood-based DNA methylation data and have demonstrated utility in predicting cardiometabolic, inflammatory, and disease-related traits in adult populations(20). While buccal DNA methylation differs from blood-derived profiles, prior work suggests that a subset of methylation signatures may be shared across tissues, enabling the potential extension of blood-derived methylation profile scores to buccal samples(21, 23, 24). Moreover, McKinnon et al. showed that around 41% of buccal-derived EpiScores were associated with low GA at birth, including EpiScores for chemokines, growth factors, proteins involved in neurogenesis and vascular development, cell membrane proteins and receptors, and other immune proteins^17^. Although these algorithms were developed in peripheral blood, buccal epithelial cells provide a minimally invasive tissue that is well suited for neonatal and pediatric studies. EpiScores may therefore yield useful information about neonatal health as it relates to preterm birth, particularly by capturing underlying biological pathways linked to both typical developmental and pathologic (e.g. inflammation) processes.

The objective of this study was to leverage data from a large multisite cohort of infants born before 30 weeks of gestation to examine associations between DNA methylation derived EpiScores and neonatal morbidity burden. We first evaluated all 111 EpiScores in relation to cumulative neonatal multimorbidity (0-1 vs. ≥2 morbidities), a clinically meaningful phenotype of cumulative neonatal complications. Then to support cross-tissue applicability, we assessed correlations between blood- and buccal-derived EpiScores using an independent dataset with paired blood and buccal DNA methylation measurements and identified a subset of scores with moderate to strong concordance. These findings provide empirical support for the cross-tissue stability of selected EpiScores, while recognizing that this analysis does not constitute formal validation of these algorithms in neonatal buccal tissue. As secondary analyses, we analyzed the association between these EpiScores with stepwise increase in the morbidity score 0 vs. 1, 2, or 3+) and with the four individual neonatal morbidities themselves. This approach aimed to identify whether increasing morbidity burden and specific complications were associated with distinct epigenetic signatures reflecting underlying biological pathways.

## Methodology

### Study Population

The Neonatal Neurobehavior and Outcomes in Very Preterm Infants (NOVI) Study comprises a multisite cohort of very preterm infants (VPT) born between April 2014 and June 2016 and cared for in nine university-affiliated Neonatal intensive care units (NICUs) in Providence, RI, Grand Rapids, MI, Kansas City, MO, Honolulu, HI, Winston-Salem, NC, and Torrance and Long Beach CA. All NICUs were participating in the Vermont Oxford Network (VON).

Infants were eligible for the NOVI Study if they were born <30 weeks GA, had a parent able to read and speak English or Spanish, and resided within three hours of the NICU and follow-up clinic. Exclusion criteria included maternal age <18 years, maternal cognitive impairment, major congenital anomalies, maternal death, or infant death prior to NICU discharge. Enrollment occurred once survival was deemed likely by the attending neonatologist(25, 26). Written informed consent was obtained from all participating mothers, and study procedures were approved by local institutional review boards.

Maternal demographic data were collected via interview, including age, race/ethnicity, education, and socioeconomic status (SES), assessed using the Hollingshead Index (level V indicating low SES)(27). Infant clinical data were abstracted from medical records and included birthweight, gestational age, length of NICU stay, outborn status, and neonatal morbidities. Gestational age was determined using criteria established by the Extremely Low Gestational Age Newborns (ELGAN) Study, prioritizing embryo retrieval or insemination dates, followed by fetal ultrasound, last menstrual period, or neonatologist assessment. Postmenstrual age (PMA) was defined as gestational age at birth plus NICU length of stay. Buccal epithelial cells were collected at NICU discharge (±3 days) for epigenomic analyses. Of the 704 enrolled infants, buccal samples were obtained from 624, with 542 passing quality control for downstream analyses. Of these, 532 had complete data for neonatal morbidities and covariates, and were included in this analysis.

### Neonatal Morbidities

Neonatal morbidities previously validated by Bassler and colleagues (11)were defined using standardized VON (28) definitions and included BPD, ROP, culture-confirmed infection (sepsis or NEC), and severe brain injury (SBI). BPD was defined as the need for supplemental oxygen at 36 weeks PMA, regardless of severity. NEC was defined using clinical and radiographic criteria, and sepsis was defined by a positive blood culture(29). SBI was assessed using cranial ultrasound imaging performed during early postnatal life and near discharge. Ultrasounds were reviewed by trained neuroradiologists using ELGAN criteria, with abnormalities classified as present if confirmed by at least two readers. SBI included periventricular leukomalacia, or moderate to severe ventricular dilation with or without intraventricular hemorrhage(30). A cumulative neonatal morbidity risk score was calculated by summing the number of morbidities experienced (BPD, severe ROP, SBI, NEC/sepsis). Consistent with our hypothesis that neonatal multimorbidity represents a clinically meaningful phenotype, infants were classified as having 0/1 or 2+ morbidities for the primary analyses; sensitivity analyses examined alternative morbidity classifications and individual morbidities.

### DNA Methylation Measurement and Preprocessing

DNA was extracted from buccal swabs using the Isohelix system (Boca Scientific) and quantified using a Qubit fluorometer (Thermo Fisher, Waltham, MA, USA). Samples were standardized to 500 ng total DNA, bisulfite-converted using the EZ DNA Methylation Kit, and assayed using the Illumina MethylationEPIC V1 Beadarray. Samples were randomized across plates and chips to minimize batch effects.

Quality control excluded samples with >5% of probes failing detection (p > 1×10□□), sex mismatches, or incomplete covariate data. Functional normalization and beta-mixture quantile (BMIQ) normalization were applied (31). Probes on sex chromosomes, probes containing SNPs, cross-reactive probes, and probes with low variability were removed. After preprocessing, 706,278 CpG sites from 532 samples were retained for analysis. DNA methylation data are publicly available via NCBI GEO (accession GSE128821)(31–33).

### EpiScore calculation

EpiScores were calculated using the web-based MethylDetectR platform, as described by Gadd et al (20) (34). MethylDetectR implements previously developed DNA methylation-based prediction models that estimate circulating protein levels from Illumina EPIC DNA methylation data. These prediction models were trained in large adult blood-based cohorts using elastic net regression, which selects informative CpG sites and assigns regression coefficients (weights) to generate methylation-derived proxies of circulating protein concentrations. The reference algorithms included 12,484 CpG sites with corresponding coefficients across the EpiScore models. Thus, EpiScores were derived from DNA methylation data and did not require direct measurement of protein concentrations. The resulting EpiScores therefore represent externally trained DNA methylation signatures that proxy protein-related biological pathways rather than direct measurements of protein abundance.

We uploaded normalized buccal DNA methylation data generated using the Illumina Infinium MethylationEPIC BeadChip to MethylDetectR and generated 109 available EpiScores for each of the 532 infants included in this study. In addition, IL-6 and CRP EpiScores were calculated using previously published algorithms, yielding a total of 111 EpiScores (20) (Supplement table 1).

### Cross-tissue concordance: Blood-buccal EpiScore correlation

Because EpiScores were originally derived from blood-based DNA methylation data and our study relied on buccal DNA methylation data, we evaluated cross-tissue concordance using an independent, external dataset (GEO accession: GSE111165). This dataset includes paired blood and buccal samples collected from living donors with medical epilepsy, aged 5–61 years (N=21). DNA methylation was measured using the Infinium HumanMethylationEPIC BeadChip, with paired buccal and blood samples (35). We also applied the MethylDetectR method to calculate 109 EpiScores for this dataset and did separate calculations for IL6 and CRP Episcore. We performed Pearson correlation tests between EpiScores derived from blood and buccal DNA methylation data to identify EpiScores that were strongly correlated between tissues, using a threshold of *r* > 0.50 (p<0.05).

### Statistical Analysis

We first examined associations between neonatal multi-morbidity (0/1 morbidity vs 2+ morbidities) and all 111 available EpiScores derived from buccal DNA methylation. Linear regression models were fitted for each EpiScore, adjusting for sex, study site, postmenstrual age (PMA) at DNA methylation collection (gestational age at birth plus postnatal age at NICU discharge), to account for differences in developmental age at the time of biospecimen collection, epithelial cell proportions, and sample plate (Figure 1). Given that EpiScores were originally developed using blood-derived DNA methylation data and we used buccal swab collections, we leveraged additional data from an external source (35) to perform cross tissues concordance analyses using Pearson correlation between 111 EpiScores calculated using blood and buccal swabs. We also performed three sensitivity analyses. First, we evaluated the robustness of the primary findings by additionally adjusting for gestational age at birth. Second, we assessed whether associations with EpiScores exhibited stepwise increases in magnitude with increasing morbidity burden (0 vs. 1, 2, or 3+). Finally, we examined associations between each individual neonatal morbidity (BPD, severe ROP, sepsis/NEC, and severe brain injury) and individual EpiScores. All sensitivity analyses were conducted with adjusted linear regression models.

**Figure 1.**
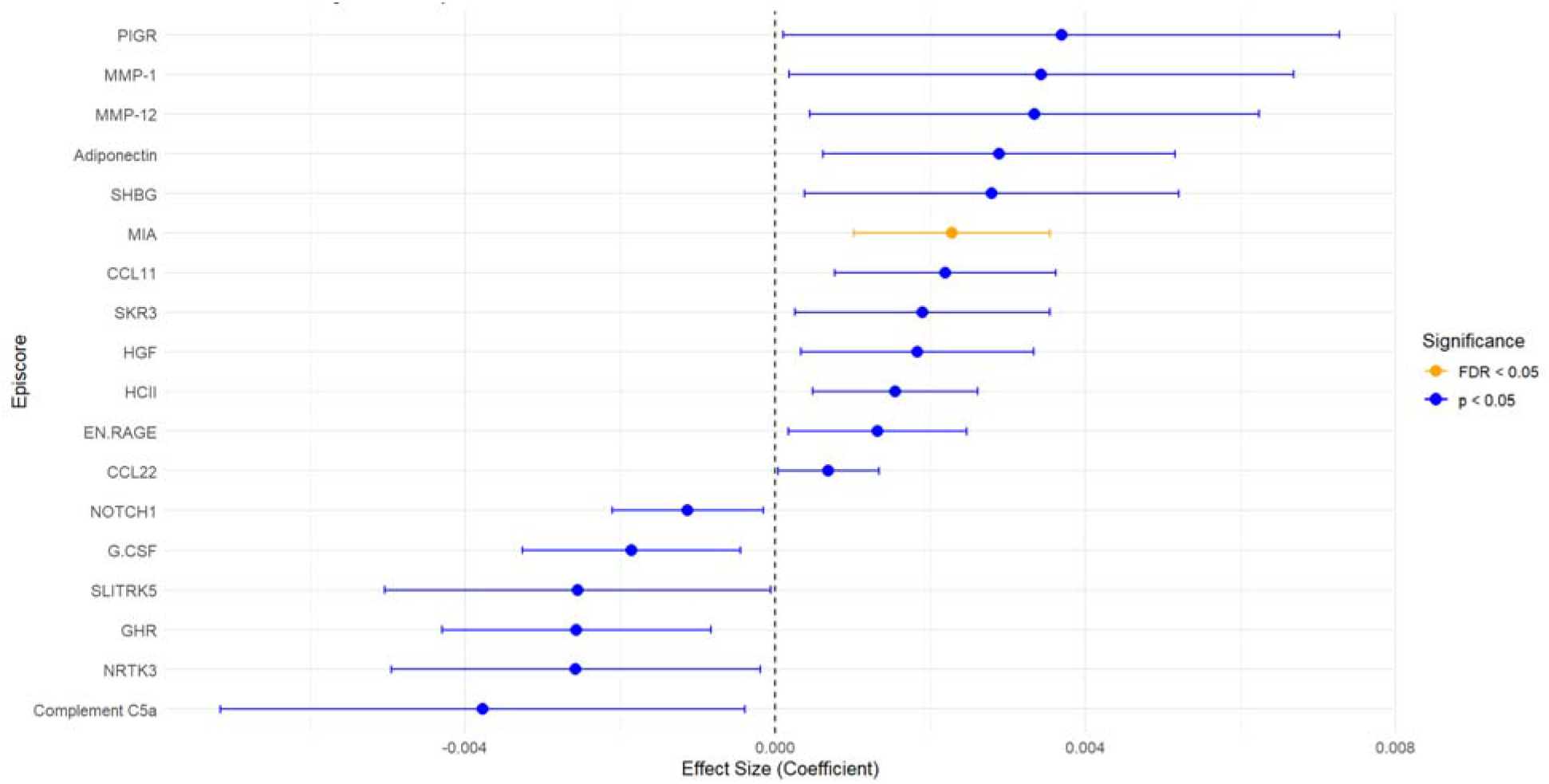
Coefficients and confidence intervals from linear regression models testing the associations between multi-morbidity (score of 2+) with 18 Episcores (adjusted for sex, study site, age at buccal sampling, epigenetic cell type, and sample plate) from full analysis of 111 EpiScores

## Results

### Demographics

Our analytical set included 537 infants from the NOVI study with DNAm data and complete neonatal morbidity risk scores. Most infants experienced at least one morbidity, with 410 (77%) having 0 or 1, and 122 (23%) having two or more complications. The mean gestational age at birth was 27.0 weeks, and mean age at buccal swab collection was 39.2 weeks post menstrual age; infants with higher risk scores (2 or more) had lower gestational age (25.6 weeks) and higher age at buccal swab collection (42.0 weeks). Our samples had more male infants 298 (55%), 72 (13%) of mothers had less than high school education, and 42 (8%) were in the lowest SES group. Among individual morbidities, BPD was most common (276, 52%), followed by infection (101, 19%), SBI (68, 13%), and severe ROP (33, 6%) (Table 1).

**Table 1.**
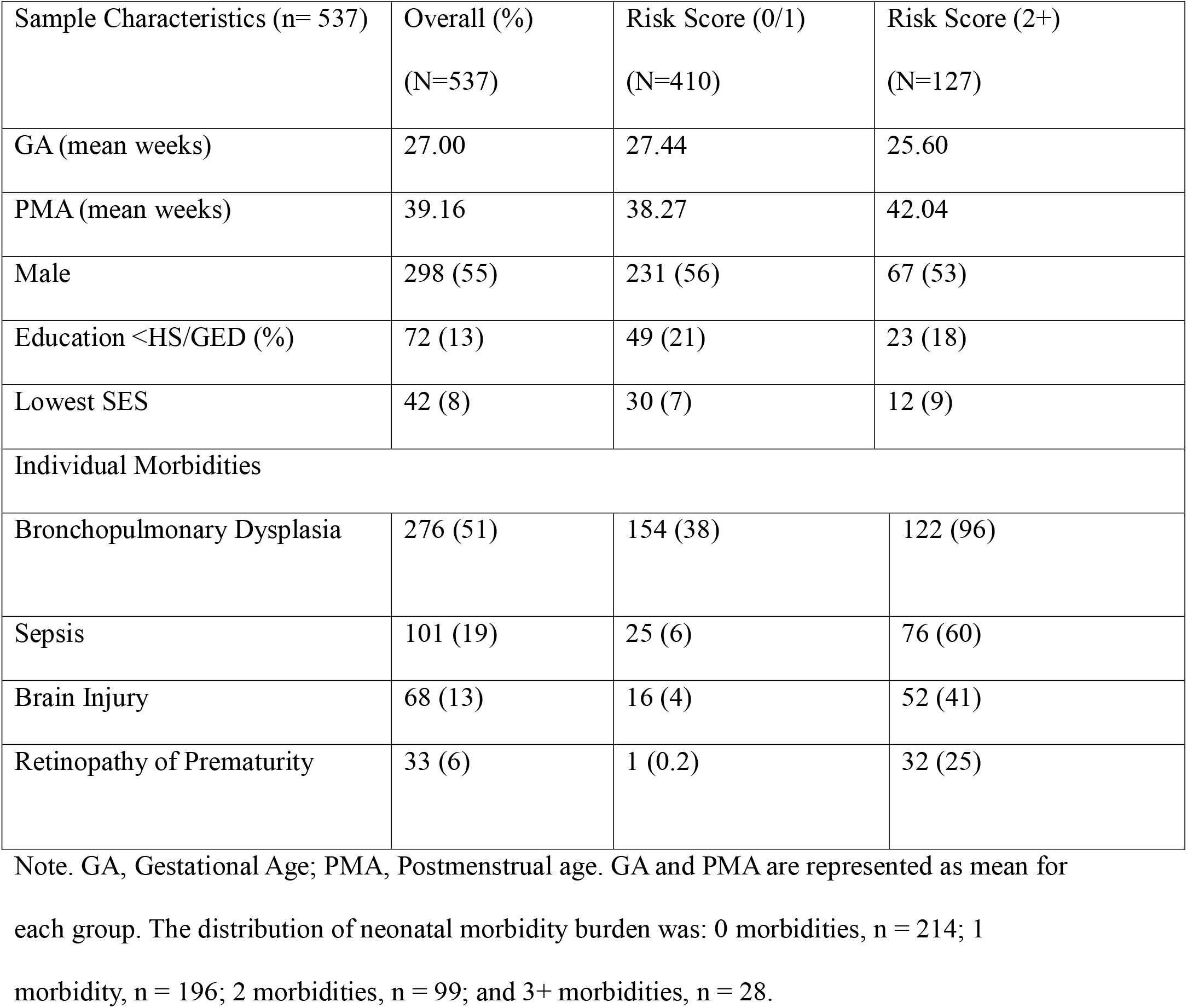
General characteristics of the study population overall and stratified by the number of morbidities.

| Sample Characteristics (n= 537) | Overall (%)<br>(N=537) | Risk Score (0/1)<br>(N=410) | Risk Score (2+)<br>(N=127) |
| --- | --- | --- | --- |
| GA (mean weeks) | 27.00 | 27.44 | 25.60 |
| PMA (mean weeks) | 39.16 | 38.27 | 42.04 |
| Male | 298 (55) | 231 (56) | 67 (53) |
| Education <HS/GED (%) | 72 (13) | 49 (21) | 23 (18) |
| Lowest SES | 42 (8) | 30 (7) | 12 (9) |
| Individual Morbidities |  |  |  |
| Bronchopulmonary Dysplasia | 276 (51) | 154 (38) | 122 (96) |
| Sepsis | 101 (19) | 25 (6) | 76 (60) |
| Brain Injury | 68 (13) | 16 (4) | 52 (41) |
| Retinopathy of Prematurity | 33 (6) | 1 (0.2) | 32 (25) |

Note. GA, Gestational Age; PMA, Postmenstrual age. GA and PMA are represented as mean for each group. The distribution of neonatal morbidity burden was: 0 morbidities, n = 214; 1 morbidity, n = 196; 2 morbidities, n = 99; and 3+ morbidities, n = 28.

### Neonatal multi-morbidity and Buccal EpiScores

Across the full set of 111 EpiScores, 18 associations were nominally statistically significant (p < 0.05); however, after correction for multiple testing using the Benjamini-Hochberg method (FDR), only one association remained statistically significant at FDR ≤ 0.05.

Higher neonatal morbidity risk was associated with higher MIA (Melanoma Inhibitory Activity) EpiScore (β = 0.0023, SE = 0.0006, p = 4.4 × 10□□, FDR = 0.048) (Figure 1).

As a sensitivity analysis, we further adjusted the primary models, which already accounted for postmenstrual age at DNA collection, by additionally including gestational age at birth to evaluate whether the observed associations were robust to alternative characterization of developmental age. Although effect estimates were modestly attenuated overall, 6 of the 18 nominally associated EpiScores remained significant (p<0.05), including MIA, which remained the strongest association (β attenuated from 0.0023 to 0.0018, p<0.05) (Supplement figure 1).

We also examined whether alternative operationalization of morbidity burden, modeled as a 4-level factor (0 morbidities vs. 1, 2, or 3+), altered our conclusions. Among the 18 identified EpiScores, infants with a single morbidity generally did not differ from those with no morbidities, whereas effect estimates for infants with 2 or 3+ morbidities were typically similar in magnitude and further from the null (Supplement figure 2). Although confidence intervals for most category-specific estimates overlapped the null, this pattern was consistent with our primary dichotomous definition of neonatal multimorbidity as a distinct phenotype of cumulative morbidity burden rather than a dose-response relationship. Only GHR demonstrated evidence of progressively larger effect estimates with increasing morbidity burden. The relatively small numbers of infants within these more granular morbidity categories limited power to distinguish differences between them.

### Cross-tissue concordance results

Of the 111 EpiScores, 35 showed moderate to strong correlations between blood and buccal samples (Pearson correlation coefficients (r) > 0.5 and p < 0.05) (Figure 2). The highest correlations were observed for Adiponectin (r = 0.86, p = 4.9 × 10□□), CHIT1 (r = 0.85, p = 1.2 × 10□□), RARRES2 (r = 0.85, p = 1.2 × 10□□), BMP1 (r = 0.84, p = 2.2 × 10□□), and Stanniocalcin-1 (r = 0.81, p = 1.1 × 10□□). Within this subset of cross-tissue concordant EpiScores, 11 showed nominal associations with neonatal morbidity including MIA and several proteins involved in inflammatory processes and neurodevelopment. For instance, higher morbidity risk was associated with higher Adiponectin (β = 0.0030, SE = 0.0012, p = 0.015), and with lower G.CSF EpiScore values (β = −0.0015, SE = 0.0007, p = 0.044).

**Figure 2.**
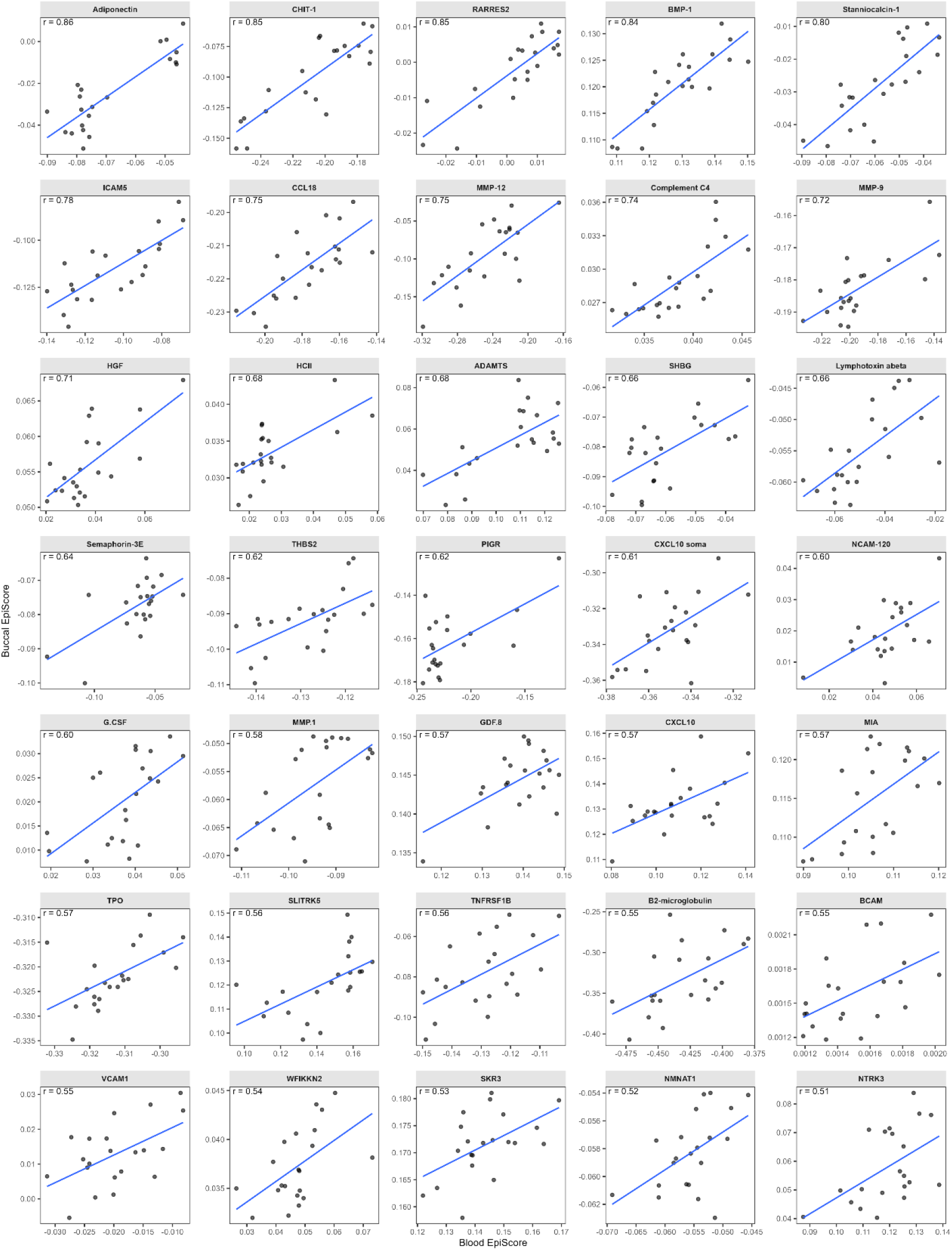
Correlation of EpiScores between blood and buccal DNA methylation, highlighting the 35 EpiScores with moderate or stronger correlations (r ≥ 0.5).

**Figure 3.**
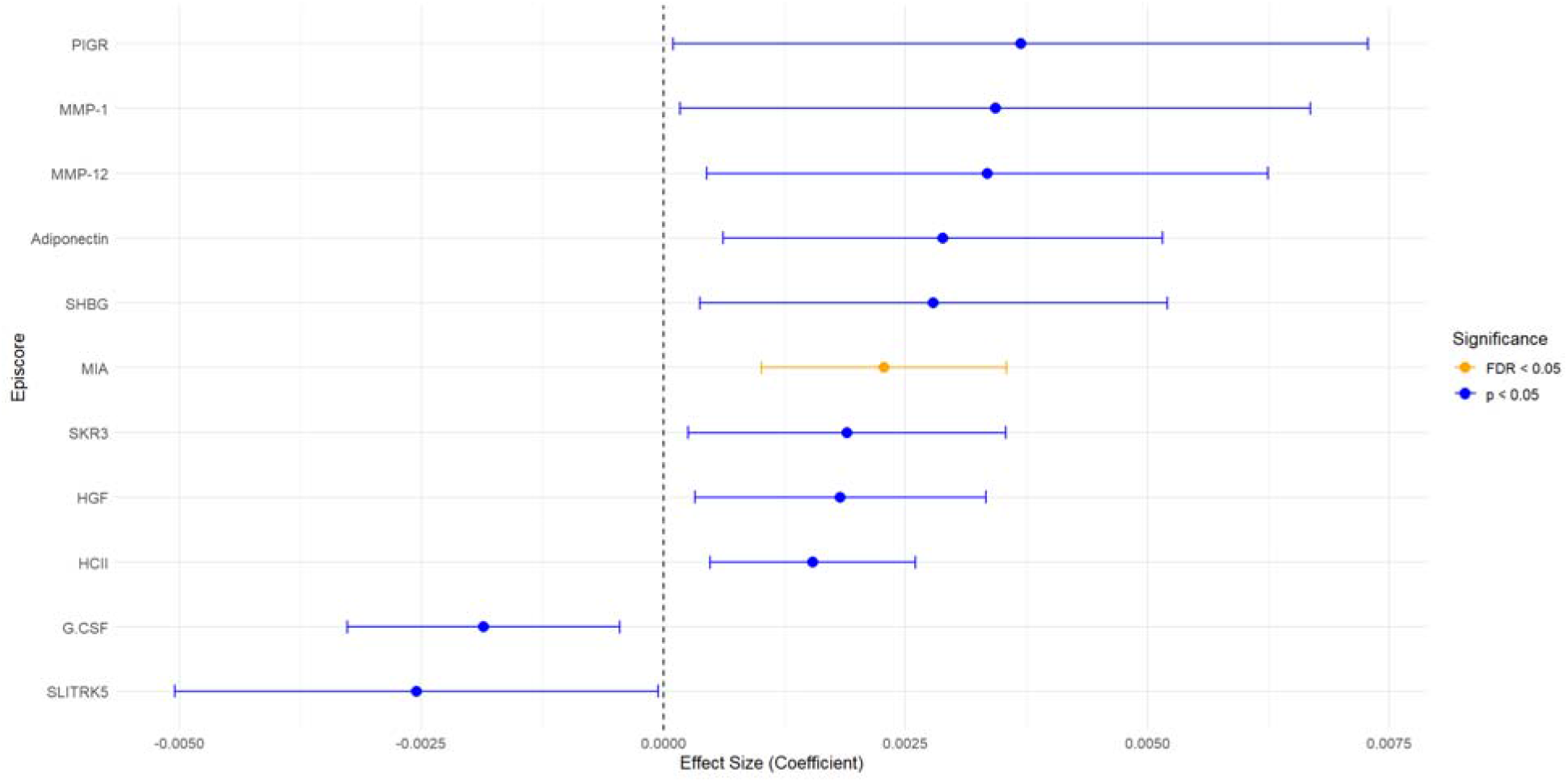
Coefficients and confidence intervals from linear regression models testing the associations between multi-morbidity (score of 2+) of the 11 morbidity-associated EpiScores among the 35 cross-tissue concordant EpiScores. (adjusted for sex, study site, age at buccal sampling, epigenetic cell type, and sample plate)

### Individual Morbidities and Buccal EpiScores

While our hypothesis focused on the impact of multimorbidity, we also explored whether individual morbidities had specific associations with EpiScores.

### Brain injury

Among infants with evidence of brain injury, 3 EpiScores were nominally associated (p < 0.05), with no associations remaining significant after FDR correction. Brain injury was associated with lower CRTAM (β = −0.0029, SE = 0.0013, p = 0.029) and lower TNFRSF1B (β = −0.0027, SE = 0.0013, p = 0.040), as well as higher HGFI (β = 0.0017, SE = 0.0008, p = 0.038).

### Bronchopulmonary Dysplasia (BPD)

Infants with BPD showed 17 nominally significant associations (p < 0.05), with no EpiScores surviving FDR correction. BPD was associated with higher CCL17 (β = 0.0029, SE = 0.0012, p = 0.013) and higher Galectin-4 (β = 0.0016, SE = 0.0006, p = 0.009), as well as lower GHR (β = −0.0010, SE = 0.0003, p = 0.0017) and lower CLEC11A e1 (β = −0.0018, SE = 0.0006, p = 0.0042).

### Necrotizing enterocolitis or sepsis

Infants with NEC or sepsis demonstrated 11 nominally significant associations (p < 0.05), with no associations significant after FDR correction. NEC/sepsis was associated with higher MIA (β = 0.0019, SE = 0.0006, p = 0.0032) and higher RARRES2 (β = 0.0028, SE = 0.0011, p = 0.010), as well as lower CHIT-1 (β = −0.0023, SE = 0.0008, p = 0.0044) and lower Aminoacylase-1 (β = −0.0038, SE = 0.0018, p = 0.034).

### Severe retinopathy of prematurity

Severe ROP was associated with 23 EpiScores at the nominal level (p < 0.05), including 1 association that remained significant after FDR correction. The GHR EpiScore remained significant after FDR correction and was lower in infants with ROP (β = −0.0054, SE = 0.0014, p = 1.33 × 10□□). At the nominal level, ROP was also associated with higher HGF (β = 0.0037, SE = 0.0012, p = 0.0024) and higher B2-microglobulin (β = 0.0102, SE = 0.0043, p = 0.019), as well as lower THBS2 (β = −0.0044, SE = 0.0016, p = 0.0053).

## Discussion

In this study, we evaluated whether EpiScores, epigenetic proxies of protein expression, were associated with neonatal morbidity burden and individual morbidities among infants who were born very preterm. Using an independent external dataset with paired blood and buccal tissues, we also show that a subset of EpiScores exhibit moderate to strong correlations between blood and buccal tissues, supporting the feasibility of extending selected blood-based methylation scores to buccal samples. Of the 111 EpiScores examined, only the MIA EpiScore was statistically significant after correction for FDR. While several additional EpiScores demonstrated nominal associations with multimorbidity and mapped to biologically plausible inflammatory, immune, metabolic, and growth-related pathways, these findings should be considered exploratory and require replication in independent cohorts.

The MIA EpiScore, which demonstrated the strongest positive association with multimorbidity, is the Melanoma-derived growth regulatory protein which is involved in growth factor regulation. MIA has been shown to inhibit the proliferation of melanoma cells(36). While MIA has been primarily studied in the context of cancer biology, its role in fetal or neonatal development remains poorly characterized. The observed association between MIA EpiScores and higher morbidity burden may reflect altered growth or tissue remodeling pathways in infants experiencing multiple neonatal complications. Further work is needed to clarify the biological relevance of this marker in early-life development and disease.

We also showed that 35 EpiScores exhibited moderate-to-strong correlations when derived from paired blood or buccal tissue samples, including MIA. These EpiScores may be particularly relevant for future and ongoing research that utilize buccal tissues for DNA methylation profiling. While no other EpiScore associations with multi-morbidity remained significant after FDR correction, it is noteworthy that 11 of the 35 cross-tissue concordant EpiScores (31%) did have nominal associations with multimorbidity. Collectively, these findings may reflect subtle but widespread alterations in protein-related biological activity associated with preterm multimorbidity, including several proteins such as adiponectin (ADPN) and granulocyte-colony stimulating factor (G.CSF) involved in pathways with strong biological relevance to the inflammatory and developmental complications commonly experienced by very preterm infants.

ADPN EpiScore showed a positive association with neonatal morbidity burden. Adiponectin is a key metabolic and immunomodulatory hormone with well-established anti-inflammatory and insulin-sensitizing properties (37). ADPN has been shown to reduce inflammatory markers such as C-reactive protein (CRP), interleukin-6 (IL-6), and tumor necrosis factor alpha (TNF-α), while elevated inflammatory signaling can suppress adiponectin production, suggesting a bidirectional regulatory relationship(38). In addition, adiponectin plays an important role in insulin sensitivity, and impaired insulin signaling has been linked to adverse neurodevelopmental and cognitive outcomes(39). In the context of very preterm infants, altered adiponectin-related epigenetic signatures may reflect early disruptions in metabolic and inflammatory regulation among those with increased morbidity burden.

G-CSF EpiScore was decreased among those with multimorbidity. G-CSF is a hematopoietic growth factor that regulates the proliferation and differentiation of hematopoietic and neural progenitor cells(40, 41). Beyond its well-known role in the immune system, research suggests that G-CSF and its receptor (G-CSFR) are expressed in the brain, indicating an important role in neuroprotection and neural repair(42). Experimental studies in animal models have demonstrated broad neuronal expression of G-CSF and G-CSFR throughout the brain, with particularly high levels in regions critical for learning, memory, and neurogenesis(42). Studies have further shown that G-CSF administration confers neuroprotective(43) and neurodegenerative(42) effects in multiple models of neurological injury and disease, including stroke, Parkinson’s disease(44), and Alzheimer’s disease(45). These findings position G-CSF as both a neurotrophic and neuroprotective factor that supports neuronal survival, reduces inflammation, and promotes functional recovery. In the context of very preterm infants, alterations in G-CSF-related epigenetic signatures may reflect early-life immune and neurodevelopmental processes that are responsive to neonatal stress, inflammation, and injury. The observed association between buccal G-CSF EpiScores and neonatal morbidity suggests that this pathway may be involved in the biological response to prematurity-related complications and further investigation in relation to later neurodevelopmental outcomes.

Thus, while the observed association with MIA represented the most statistically robust finding from our analyses, several additional EpiScores, including G-CSF and adiponectin, also demonstrated suggestive associations with multimorbidity that are biologically plausible given their known roles in inflammatory signaling, immune regulation, and neuronal health. Together, these findings support the possibility that neonatal multimorbidity in very preterm infants is accompanied by coordinated, though often subtle, dysregulation across multiple biologically relevant processes. Additionally, in our exploratory analyses of the individual neonatal morbidities, specific EpiScores showed associations that correspond with established biological mechanisms relevant to these complications.

Brain injury was associated with lower CRTAM and higher HGFI. CRTAM (Cytotoxic and Regulatory T-cell Molecule) is a member of the nectin family that plays an important role in immune regulation. Interaction of CRTAM with CADM1 promotes natural killer (NK) cell cytotoxicity and interferon-gamma (IFN-γ) secretion by CD8+ T cells and has also been implicated in NK cell–mediated tumor rejection through interaction with CADM3(46). HGFI corresponds to hepatocyte growth factor (HGF), a multifunctional growth factor that promotes cell proliferation, tissue repair, and regeneration through activation of the MET receptor tyrosine kinase pathway(47). HGF-MET signaling plays a role in brain development and neural repair, supporting neuronal survival, synaptogenesis, and neurodevelopmental processes, and has been implicated in neuroprotection and the pathophysiology of neurodevelopmental disorders(48).

Bronchopulmonary dysplasia (BPD) is one of the most prevalent morbidities in very preterm infants and was most strongly associated with lower NT-3 growth factor receptor (NTRK3) and lower Growth Hormone Receptor (GHR). EpiScore NTRK3 is a receptor tyrosine kinase that mediates the effects of neurotrophin-3 (NTF3) and plays a critical role in nervous system development and potentially cardiac development(49). Experimental models have shown that hyperoxia, similar to what neonates with BPD may experience with mechanical ventilation, can repress GHR expression, with potential acute and long-term consequences for respiratory function(50).

NEC/sepsis was negatively associated with FCRL2 EpiScores. FCRL2 (Fc receptor-like protein 2) is a CD molecule primarily expressed in B cells and is thought to play a regulatory role in normal and neoplastic B-cell development(51).

Severe ROP was associated with lower GHR EpiScores, which was also downregulated with BPD, and GHR was the only individual morbidity association that remained significant after FDR correction. GHR is a receptor for pituitary growth hormone and belongs to the type I cytokine receptor family; it plays a central role in postnatal growth and development largely through stimulating the production of IGF-1(52), and low circulating levels of IGF-1 are linked to ROP(53). Of note, MIA was positively associated with both of BPD and NEC/Sepsis, which may explain its strong association with multimorbidity in the primary analyses.

These results indicate that EpiScores derived from buccal DNA methylation capture biologically meaningful pathways including immune activation, metabolic regulation, and growth factor signaling that are relevant to overall morbidity burden in children born very preterm. Because buccal DNA methylation was measured at NICU discharge after neonatal morbidities had already occurred, these analyses cannot establish whether altered EpiScores preceded morbidity onset, contributed to susceptibility, or instead reflect the biological consequences of these complications, inflammation, intensive care treatments, or other NICU-related exposures. Therefore, the EpiScores identified here are best interpreted as biomarkers of cumulative neonatal morbidity burden rather than evidence of causal drivers of pathogenesis. Even so, these biomarkers remain valuable because they capture the integrated biological response to complex neonatal complications and may provide insight into the molecular pathways associated with overall morbidity burden and subsequent health outcomes.

This study should be interpreted while considering its limitations. Although we evaluated cross-tissue concordance using an independent dataset with paired blood and buccal DNA methylation profiles, several limitations should be considered when interpreting these findings. The cross-tissue dataset was small (n=21), consisted of individuals with epilepsy rather than healthy neonates, and included participants spanning a broad age range. Because DNA methylation patterns are influenced by tissue type, age, and disease status, the performance of blood-derived EpiScores in neonatal buccal samples may differ from that observed in this external dataset. Consequently, the observed blood-buccal concordance should be interpreted as evidence that a subset of EpiScores may generalize across tissues and ages rather than as definitive validation in neonatal populations. Nevertheless, demonstrating cross-tissue concordance represents an important strength of this study by providing empirical support for interpreting selected buccal-derived EpiScores in studies where blood collection is not feasible. Second, although we evaluated a large number of EpiScores, most associations observed in this study were nominally significant and did not remain significant after correction for multiple testing. This raises the possibility of false positive findings and suggests that results, particularly for individual morbidities, should be interpreted cautiously and considered exploratory until replicated in independent cohorts. Third, due to the cross-sectional nature of the data, we are unable to comment if EpiScores reflect the consequences of neonatal morbidity or underlying biochemical dysregulation that lead to the onset of morbid conditions in this period of development. Lastly, despite adjustment for several important covariates, residual confounding by unmeasured clinical or environmental factors, such as, treatment exposures, or NICU-specific practices, cannot be excluded.

## Conclusion

In this study, we demonstrate that DNA methylation-derived EpiScores from buccal samples can capture biologically relevant signals associated with neonatal morbidity burden and individual complications among infants born very preterm. While only the MIA EpiScore remained significant after multiple testing correction for overall morbidity burden, several nominal associations with important biological relevance were also identified. Additionally, we observed associations with individual morbidities, highlighting distinct patterns linked to immune, inflammatory, and growth-related pathways. Severe ROP showed the strongest signal, including one EpiScore (GHR) that remained significant after FDR correction, suggesting a more robust epigenetic signature for this condition. Future studies are needed to replicate these associations, examine associations with longitudinal outcomes, and further investigate the functional significance of key epigenetic markers in early development.

## Data Availability

All data produced in the present study are available upon reasonable request to the authors

## List of abbreviations

ADPN: Adiponectin
BPD: Bronchopulmonary Dysplasia
BMIQ: Beta-Mixture Quantile Normalization
CADM1: Cell Adhesion Molecule 1
CADM3: Cell Adhesion Molecule 3
CHIT1: Chitotriosidase-1
CpG: Cytosine-phosphate-Guanine
CRP: C-Reactive Protein
CRTAM: Cytotoxic and Regulatory T-cell Molecule
DNAm: DNA Methylation
ELGAN: Extremely Low Gestational Age Newborns
FCRL2: Fc Receptor-Like Protein 2
FDR: False Discovery Rate
GA: Gestational Age
G-CSF: Granulocyte Colony-Stimulating Factor
G-CSFR: Granulocyte Colony-Stimulating Factor Receptor
GEO: Gene Expression Omnibus
GHR: Growth Hormone Receptor
HGF: Hepatocyte Growth Factor
HGFI: Hepatocyte Growth Factor-Related EpiScore
IFN-γ: Interferon Gamma
IGF-1: Insulin-Like Growth Factor 1
IL-6: Interleukin-6
NEC: Necrotizing Enterocolitis
NICU: Neonatal Intensive Care Unit
NK: Natural Killer
NOVI: Neonatal Neurobehavior and Outcomes in Very Preterm Infants
NTF3: Neurotrophin-3
NTRK3: Neurotrophic Receptor Tyrosine Kinase 3
PMA: Postmenstrual Age
RARRES2: Retinoic Acid Receptor Responder Protein 2
ROP: Retinopathy of Prematurity
SBI: Severe Brain Injury
SES: Socioeconomic Status
SNP: Single Nucleotide Polymorphism
TNF-α: Tumor Necrosis Factor Alpha
TNFRSF1B: Tumor Necrosis Factor Receptor Superfamily Member 1B
VON: Vermont Oxford Network
VPT: Very Preterm

## Declarations

### Ethics approval and consent to participate

All recruitment, enrollment, and data collection procedures were approved by local institutional review boards. The primary caregiver provided written informed consent for themselves and their infants’ participation.

### Consent of Publication

Not applicable

### Availability of data and materials

Data are accessible through NCBI Gene Expression Omnibus (GEO) via accession series <u>GSE128821</u>. Statistical code for all analyses is available upon request from the first author.

### Author contributions

PP: conceptualization; methodology; funding acquisition; formal analysis; interpretation of findings; visualization of findings; preparation of the original manuscript. TME, BML, CJM: conceptualization; methodology; supervision; funding acquisition; investigation; interpretation of findings; review and editing of the manuscript. SZ, NS: interpretation of findings; review and editing of the manuscript. MC,BSC, JC, JBH, JAH, ECM, CRN, SLP, LMS, MOS: investigation; supervision; interpretation of findings; review and editing of the manuscript. TME: conceptualization; methodology; supervision; funding acquisition; investigation; data curation; interpretation of findings; review and editing of the manuscript. All authors read, edited, and approved the final manuscript.

## Acknowledgements

We would like to thank the Emory & WIH lab teams, NOVI Study Coordinators, and the NOVI families who made this work possible.

## References

1. Jensen EA, Edwards EM, Greenberg LT, Soll RF, Ehret DEY, Horbar JD. Severity of Bronchopulmonary Dysplasia Among Very Preterm Infants in the United States. Pediatrics. 2021;148(1).

2. Lekha T, Balakrishnan D, Giridhar A, Alex D, Goyal A. Retinopathy of Prematurity in Extreme Preterm and Extreme Low-birth-weight Infants: Incidence, Course, and Risk Factors. Middle East Afr J Ophthalmol. 2023;30(3):136–40.

3. Gregory KE, Deforge CE, Natale KM, Phillips M, Van Marter LJ. Necrotizing enterocolitis in the premature infant: neonatal nursing assessment, disease pathogenesis, and clinical presentation. Adv Neonatal Care. 2011;11(3):155–64; quiz 65-6.

4. Flannery DD, Edwards EM, Puopolo KM, Horbar JD. Early-Onset Sepsis Among Very Preterm Infants. Pediatrics. 2021;148(4).

5. Kidokoro H, Anderson PJ, Doyle LW, Woodward LJ, Neil JJ, Inder TE. Brain injury and altered brain growth in preterm infants: predictors and prognosis. Pediatrics. 2014;134(2):e444–53.

6. Donlon J, Hawkins K, Bhat V, Hunter K, Kushnir A, Bhandari V. Impact of bronchopulmonary dysplasia, brain injury, necrotizing enterocolitis, retinopathy of prematurity and sepsis on neurodevelopmental outcomes in premature infants. J Perinatol. 2025.

7. Alonso-Lopez P, Arroyas M, Beato M, Ruiz-Gonzalez S, Olabarrieta I, Garcia-Garcia ML. Respiratory, cardio-metabolic and neurodevelopmental long-term outcomes of moderate to late preterm birth: not just a near term-population. A follow-up study. Front Med (Lausanne). 2024;11:1381118.

8. Everson TM, O’Shea TM, Burt A, Hermetz K, Carter BS, Helderman J, et al. Serious neonatal morbidities are associated with differences in DNA methylation among very preterm infants. Clin Epigenetics. 2020;12(1):151.

9. McGowan EC, Hofheimer JA, O’Shea TM, Kilbride H, Carter BS, Check J, et al. Analysis of Neonatal Neurobehavior and Developmental Outcomes Among Preterm Infants. JAMA Netw Open. 2022;5(7):e2222249.

10. van Hasselt TJ, Dorner RA, Katheria A, Battersby C, Gale C, Lo DKH, et al. Neonatal Morbidities and Hospitalization in the First 2 Years of Life Among Infants Born Very Preterm. JAMA Netw Open. 2025;8(9):e2530123.

11. Bassler D, Stoll BJ, Schmidt B, Asztalos EV, Roberts RS, Robertson CM, et al. Using a count of neonatal morbidities to predict poor outcome in extremely low birth weight infants: added role of neonatal infection. Pediatrics. 2009;123(1):313–8.

12. Dorner RA, Li L, DeMauro SB, Schmidt B, Zangeneh SZ, Vaucher Y, et al. Association of a Count of Inpatient Morbidities with 2-Year Outcomes among Infants Born Extremely Preterm. J Pediatr. 2025;278:114428.

13. Farooqi A, Hagglof B, Sedin G, Serenius F. Impact at age 11 years of major neonatal morbidities in children born extremely preterm. Pediatrics. 2011;127(5):e1247–57.

14. Schmidt B, Roberts RS, Davis PG, Doyle LW, Asztalos EV, Opie G, et al. Prediction of Late Death or Disability at Age 5 Years Using a Count of 3 Neonatal Morbidities in Very Low Birth Weight Infants. J Pediatr. 2015;167(5):982–6 e2.

15. Vulcanescu A, Siminel MA, Dijmarescu AL, Manolea MM, Sandulescu SM, Radulescu VM, et al. Molecular Mechanisms Underlying Inflammation in Early-Onset Neonatal Sepsis: A Systematic Review of Human Studies. J Clin Med. 2025;14(15).

16. Chen CC, Lin YC, Lee CY, Kuo CC, Chang TH, Huang CC. Proinflammatory state and metabolic dysregulation linking delayed feeding progression to extrauterine restricted head growth in extremely preterm infants. BMC Med. 2025;23(1):701.

17. Seassau A, Munos P, Gire C, Tosello B, Carchon I. Neonatal Care Unit Interventions on Preterm Development. Children (Basel). 2023;10(6).

18. Litt JS, Belfort MB, Everson TM, Haneuse S, Tiemeier H. Neonatal multimorbidity and the phenotype of premature aging in preterm infants. Pediatr Res. 2025;97(7):2258–66.

19. Paniagua U, Lester BM, Marsit CJ, Camerota M, Carter BS, Check JF, et al. Epigenetic age acceleration, neonatal morbidities, and neurobehavioral profiles in infants born very preterm. Epigenetics. 2023;18(1):2280738.

20. Gadd DA, Hillary RF, McCartney DL, Zaghlool SB, Stevenson AJ, Cheng Y, et al. Epigenetic scores for the circulating proteome as tools for disease prediction. Elife. 2022;11.

21. Smikle R, McKinnon K, Vaher K, Turner H, Cruickshank H, Amir R, et al. Protein epigenetic scores derived in neonatal saliva as biomarkers of childhood cognition. Mol Psychiatry. 2026.

22. Winchester P, Nilsson E, Beck D, Skinner MK. Preterm birth buccal cell epigenetic biomarkers to facilitate preventative medicine. Sci Rep. 2022;12(1):3361.

23. McKinnon K, Conole ELS, Vaher K, Hillary RF, Gadd DA, Binkowska J, et al. Epigenetic scores derived in saliva are associated with gestational age at birth. Clin Epigenetics. 2024;16(1):84.

24. Zarandooz S, Raffington L. Applying blood-derived epigenetic algorithms to saliva: cross-tissue similarity of DNA-methylation indices of aging, physiology, and cognition. Clin Epigenetics. 2025;17(1):61.

25. O’Shea TM, Allred EN, Dammann O, Hirtz D, Kuban KC, Paneth N, et al. The ELGAN study of the brain and related disorders in extremely low gestational age newborns. Early Hum Dev. 2009;85(11):719–25.

26. McElrath TF, Hecht JL, Dammann O, Boggess K, Onderdonk A, Markenson G, et al. Pregnancy disorders that lead to delivery before the 28th week of gestation: an epidemiologic approach to classification. Am J Epidemiol. 2008;168(9):980–9.

27. AB H. Four factor index of social status. 1975:47–55.

28. Edwards EM, Ehret DEY, Soll RF, Horbar JD. Vermont Oxford Network: a worldwide learning community. Transl Pediatr. 2019;8(3):182–92.

29. Network VO. Manual of operations: Part 2. Data definitions and infant data forms. Burlington, VT: Vermont Oxford Network. 2018.

30. Kuban K, Adler I, Allred EN, Batton D, Bezinque S, Betz BW, et al. Observer variability assessing US scans of the preterm brain: the ELGAN study. Pediatr Radiol. 2007;37(12):1201–8.

31. Teschendorff AE, Marabita F, Lechner M, Bartlett T, Tegner J, Gomez-Cabrero D, et al. A beta-mixture quantile normalization method for correcting probe design bias in Illumina Infinium 450 k DNA methylation data. Bioinformatics. 2013;29(2):189–96.

32. Pidsley R, Zotenko E, Peters TJ, Lawrence MG, Risbridger GP, Molloy P, et al. Critical evaluation of the Illumina MethylationEPIC BeadChip microarray for whole-genome DNA methylation profiling. Genome Biol. 2016;17(1):208.

33. Logue MW, Smith AK, Wolf EJ, Maniates H, Stone A, Schichman SA, et al. The correlation of methylation levels measured using Illumina 450K and EPIC BeadChips in blood samples. Epigenomics. 2017;9(11):1363–71.

34. Hillary RF, Marioni RE. MethylDetectR: a software for methylation-based health profiling. Wellcome Open Res. 2020;5:283.

35. Braun PR, Han S, Hing B, Nagahama Y, Gaul LN, Heinzman JT, et al. Genome-wide DNA methylation comparison between live human brain and peripheral tissues within individuals. Transl Psychiatry. 2019;9(1):47.

36. Kolanczyk M, Mautner V, Kossler N, Nguyen R, Kuhnisch J, Zemojtel T, et al. MIA is a potential biomarker for tumour load in neurofibromatosis type 1. BMC Med. 2011;9:82.

37. Nguyen TMD. Adiponectin: Role in Physiology and Pathophysiology. Int J Prev Med. 2020;11:136.

38. Ouchi N, Walsh K. Adiponectin as an anti-inflammatory factor. Clin Chim Acta. 2007;380(1-2):24–30.

39. Rizzo MR, Fasano R, Paolisso G. Adiponectin and Cognitive Decline. Int J Mol Sci. 2020;21(6).

40. Schabitz WR, Schneider A. New targets for established proteins: exploring G-CSF for the treatment of stroke. Trends Pharmacol Sci. 2007;28(4):157–61.

41. Solaroglu I, Jadhav V, Zhang JH. Neuroprotective effect of granulocyte-colony stimulating factor. Front Biosci. 2007;12:712–24.

42. Schneider A, Kruger C, Steigleder T, Weber D, Pitzer C, Laage R, et al. The hematopoietic factor G-CSF is a neuronal ligand that counteracts programmed cell death and drives neurogenesis. J Clin Invest. 2005;115(8):2083–98.

43. Schabitz WR, Kollmar R, Schwaninger M, Juettler E, Bardutzky J, Scholzke MN, et al. Neuroprotective effect of granulocyte colony-stimulating factor after focal cerebral ischemia. Stroke. 2003;34(3):745–51.

44. Meuer K, Pitzer C, Teismann P, Kruger C, Goricke B, Laage R, et al. Granulocyte-colony stimulating factor is neuroprotective in a model of Parkinson’s disease. J Neurochem. 2006;97(3):675–86.

45. Tsai KJ, Tsai YC, Shen CK. G-CSF rescues the memory impairment of animal models of Alzheimer’s disease. J Exp Med. 2007;204(6):1273–80.

46. Arase N, Takeuchi A, Unno M, Hirano S, Yokosuka T, Arase H, et al. Heterotypic interaction of CRTAM with Necl2 induces cell adhesion on activated NK cells and CD8+ T cells. Int Immunol. 2005;17(9):1227–37.

47. Xiao GH, Jeffers M, Bellacosa A, Mitsuuchi Y, Vande Woude GF, Testa JR. Anti-apoptotic signaling by hepatocyte growth factor/Met via the phosphatidylinositol 3-kinase/Akt and mitogen-activated protein kinase pathways. Proc Natl Acad Sci U S A. 2001;98(1):247–52.

48. Desole C, Gallo S, Vitacolonna A, Montarolo F, Bertolotto A, Vivien D, et al. HGF and MET: From Brain Development to Neurological Disorders. Front Cell Dev Biol. 2021;9:683609.

49. Chalazonitis A. Neurotrophin-3 as an essential signal for the developing nervous system. Mol Neurobiol. 1996;12(1):39–53.

50. Vohlen C, Mohr J, Fomenko A, Kuiper-Makris C, Grzembke T, Aydogmus R, et al. Dynamic Regulation of GH-IGF1 Signaling in Injury and Recovery in Hyperoxia-Induced Neonatal Lung Injury. Cells. 2021;10(11).

51. Nuckel H, Collins CH, Frey UH, Sellmann L, Durig J, Siffert W, et al. FCRL2 mRNA expression is inversely associated with clinical progression in chronic lymphocytic leukemia. Eur J Haematol. 2009;83(6):541–9.

52. Dehkhoda F, Lee CMM, Medina J, Brooks AJ. The Growth Hormone Receptor: Mechanism of Receptor Activation, Cell Signaling, and Physiological Aspects. Front Endocrinol (Lausanne). 2018;9:35.

53. Liegl R, Lofqvist C, Hellstrom A, Smith LE. IGF-1 in retinopathy of prematurity, a CNS neurovascular disease. Early Hum Dev. 2016;102:13–9.

