## supplement figure 1 for "Buccal DNA Methylation-Derived EpiScores are Associated with Neonatal Morbidity Burden in Infants born Very Preterm"

Supplement figure 1. Sensitivity Analysis of EpiScore Associations with Neonatal Multimorbidity After Additional Adjustment for Gestational Age at Birth

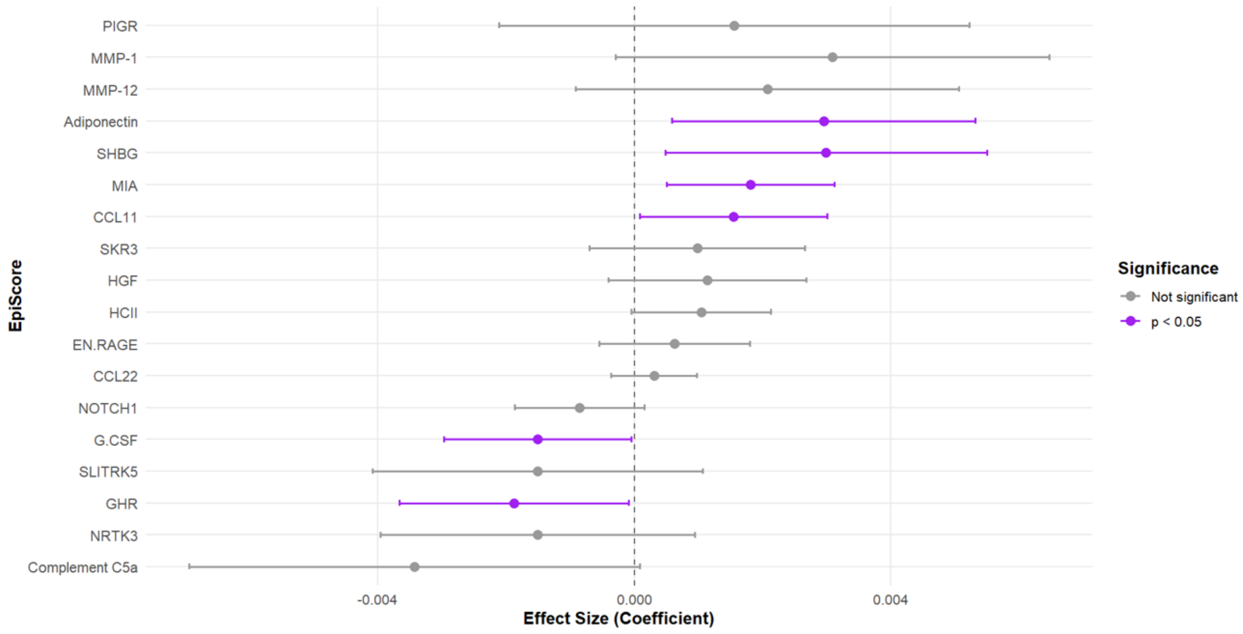
