## supplement figure 2 for "Buccal DNA Methylation-Derived EpiScores are Associated with Neonatal Morbidity Burden in Infants born Very Preterm"

Supplement Figure 2. Associations between EpiScores and increasing neonatal morbidity burden.

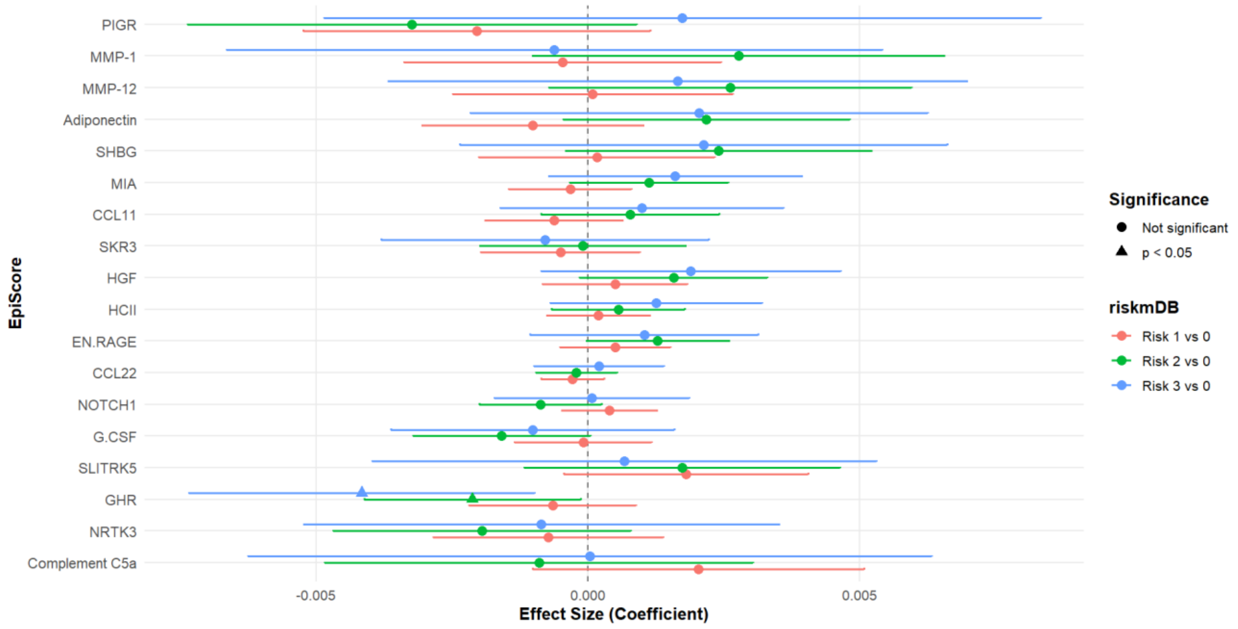


Forest plot showing adjusted associations between neonatal morbidity burden and the 18 EpiScores that were nominally associated with multimorbidity in the primary analysis. Infants with zero morbidities served as the reference group, with effect estimates shown separately for infants with 1, 2, and 3 morbidities. Points represent regression coefficients and horizontal lines represent 95% confidence intervals. Models were adjusted for sex, study site, postmenstrual age at buccal sample collection, epithelial cell proportion, and sample plate. Triangles indicate nominally significant associations ($p < 0.05$).
